# Endovascular structures of the basilar artery: forms of the basilar nonfusion spectrum

**DOI:** 10.1101/2025.01.07.25320164

**Authors:** Mikołaj Sługocki, Radosław Rzepliński, Sylwia Tarka, Tomasz Stępień, Michał Tomaszewski, Michał Kucewicz, Gabriela Kępczyńska, Jerzy Małachowski, Bogdan Ciszek

**Author notes:** Corresponding author: Radosław Rzepliński, 5 Chałubińskiego Street, 02-004 Warsaw, Poland.

## Abstract

**Introduction:** Cerebrovascular diseases are a growing social and clinical problem, and their pathogenesis is currently being thoroughly investigated. The intravascular anatomy of the cerebral circulation remains poorly understood, although an increasing number of endovascular interventions are being conducted during the course of diagnosis and treatment. The purpose of this study was to describe intravascular structures in the vertebrobasilar system and to investigate the hemodynamic consequences of their presence.

**Methods:** Thirty anatomical specimens of the human brain were analyzed via angioscopy of the vertebrobasilar system, and the presence of intravascular structures was documented. Additional histological studies were performed. The effect on blood flow was simulated using computational fluid dynamics by studying 5 different cases.

**Results:** In 8 cases (26.7%), the following endovascular structures were visualized: 6 strings, one septum, and one chord. The histological structure showed a layered pattern, resembling that of the arterial wall: the outermost tunica intima, the innermost adventitia and the tunica media in between. Blood flow simulations revealed several areas of disturbed flow, including areas of low wall shear stress and recirculations, as well as areas of elevated wall shear stress.

**Conclusions:** Intravascular structures are common in the basilar artery. The reason for their formation is the incomplete fusion of the longitudinal neural arteries, and together with fenestrations, they belong to the basilar nonfusion spectrum. The presence of structures can cause technical difficulties and ischemic complications related to endovascular interventions. Hemodynamic changes caused by endovascular structures can promote atherosclerosis, thromboembolism, narrowing of the pontine arteries and the development of aneurysms.

## 1. Introduction

In recent decades, the approach to the treatment of various cerebrovascular diseases (e.g., intracranial aneurysms, subarachnoid hemorrhages, and ischemic strokes) has significantly changed due to the dynamic development of endovascular techniques.^1-4^ Less invasive interventional methods have since attained a well-established position in the medical arsenal against cerebrovascular conditions. The progress does not seem to decelerate as we witness the emergence of large new multicenter studies demonstrating the effectiveness of interventional techniques in conditions previously considered to respond better to typical treatment, e.g., recent advances in the fields of basilar thrombectomy or embolization of the middle meningeal artery in chronic subdural hematoma.^1,5^ At the foundation of interventional neuroradiology lies thorough knowledge of the cerebrovascular anatomy.^6^ While the external and radiological anatomy is well described in most aspects,^7-13^ the endovascular anatomy of cerebral arteries seems to be greatly overlooked. To date, only a few researchers have addressed this issue, mainly by means of traditional dissection and imaging.^14-18^ Conversely, endoscopy has been previously used to assess the endovascular anatomy of the coronary sinus and superior sagittal sinus, and has yielded valuable results in both cases.^19,20^

Considering all the above, the authors of this study have decided to incorporate endovascular endoscopy (angioscopy) into the study of the internal anatomy of vertebrobasilar arteries with the aim of describing intravascular structures and investigating the hemodynamic consequences using numerical fluid mechanics.

## 2. Methods

### 2.1. Study approval

The material for the study was provided by the Department of Descriptive and Clinical Anatomy at the Medical University of Warsaw. The study protocol was approved by the Ethics Committee of the Medical University of Warsaw, Poland (number 138/2023).

### 2.2. Study material and angioscopy protocol

The material examined in this work comprised 30 unfixed cadaveric cerebral specimens (4 female and 26 male donors, age range 19–77, mean age of 50 years). In each case, cerebrovascular accidents, CNS tumors and head trauma as causes of death were ruled out. The cerebra were carefully removed from the skull. Immediately after retrieval, the specimens were placed in a 4 °C freezer. The brains were subsequently examined within 72 hours of acquisition. First, each specimen was placed on a dissection table with the basal surface of the brain facing upward and carefully dissected using a OPMI Pico surgical microscope (Carl Zeiss, Germany) and microsurgical instruments. Consecutive subarachnoid cisterns (prepontine, interpeduncular and ambiens bilaterally) were opened. Subsequently, a 20G intravenous cannula was inserted into the right vertebral artery (VA) and ligated to prevent it from slipping out of the vessel. The arteries were then rinsed with 0.9% saline solution. Subsequently, microvascular clips were placed on major branches of the vertebrobasilar arterial system. Closing the lumen of these arteries and filling them with saline solution enabled the simulation of their *in vivo* size and conformation. Smaller branches were deliberately left patent to prevent rupture of the arterial wall and potential damage to intraluminal structures. Additionally, the specimens were evaluated for any signs of visible pathology.

The arteries were then examined using an endoscopy column and a rigid endoscope (1,9 mm diameter, 0°, Karl Storz, Germany), which was inserted through the left VA, advanced toward the bifurcation of the basilar arteries (BBA) and withdrawn in the same way. A small incision was then created in the ventral part of the wall of BA through which the endoscope was introduced, leading toward the union of the vertebral arteries (UVA). Saline solution was administered to the arteries throughout the duration of the endoscopic examination. The presence of atherosclerosis in the VA, BA and posterior cerebral artery (PCA) was assessed, and the severity of atherosclerosis was classified as no atherosclerosis, atheromas, fibroatheromas, or complicated lesions. In the case of intraluminal structures, the arterial segment containing them was removed and fixed in 10% buffered formalin solution for histological examination (for no less than 7 days). The endovascular structures were measured after fixation.

### 2.3. Histological studies

The specimens were subsequently processed and immersed in standard paraffin blocks, which were cut into 5 μm sections and stained with standard hematoxylin-eosin and Mallory trichrome. The histological sections were then viewed under an Olympus BX53 optical microscope at a magnification of 25–400x, and images were acquired using an Olympus UC90 camera and cellSens Dimension software (Olympus Corporation, Japan).

### 2.4. Blood flow simulation methodology

The application of numerical fluid mechanics allows for the analysis of blood flow within vascular lesions, such as strings. The flow analysis was performed using the Ansys Fluent numerical fluid mechanics solver. Owing to the absence of specific data regarding the stiffness and strength of the tissue forming the string, a nondeformable model was employed for the simulations. This approach has inherent limitations, such as a lack of tissue deformation under blood flow pressure, potentially leading to higher calculated values of the wall shear stress (WSS). For the simulations, the simplified geometry of the basilar artery (BA) was selected (Fig. 1). The diameter of the vessel was 4.8 mm.

**Figure 1.**
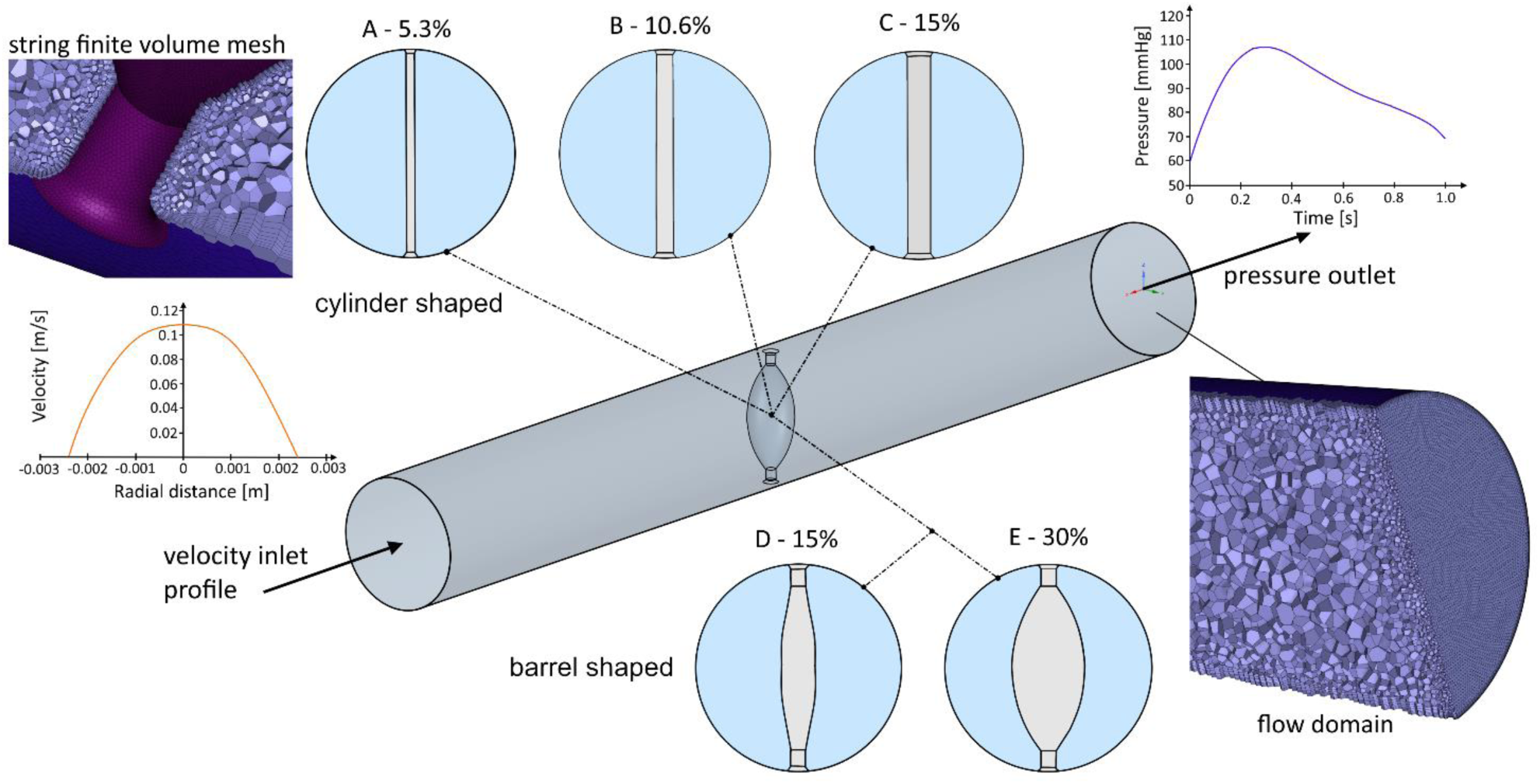
Numerical model of the basilar artery and an overview of the analyzed cases.

A two-stage flow modeling methodology was implemented. In the initial stage, a preliminary analysis was conducted to generate a full velocity profile at the inlet of the simulated vessel. On the basis of the expected flow conditions within the vessel, the characteristic dimension of the finite volume grid was subsequently set to 0.02 mm. The computational domain was discretized using Ansys Fluent Meshing. To better capture local wall phenomena, which were critical for this analysis, local mesh refinement was applied to the outer contour of the geometry. Seven boundary wall layers were utilized, with a progressive layer height increase factor of 120%. Our previous work demonstrated that the use of polyhedral elements optimizes computational efficiency while maintaining high accuracy of the results. ^21^ The maximum edge size for the finite volumes was 0.3 mm, and the minimum element quality was closely monitored, maintaining a value of 0.45. A mesh convergence study was performed during the simulation process, resulting in a domain comprising approximately 1 million elements.

The next step was to simulate blood flow in the prepared domains. Properties corresponding to blood parameters were applied for the fluid: density of 1060 kg/m^3^ and viscosity described by the Carreau model, which considers the dependence of viscosity on shear rate. ^22^ A simplification in the form of no-slip conditions on the inner walls of the vessel was applied. Time-independent (steady state) analyses with 500 iterations were performed, and a parameter convergence of 10^-6^ of residual values was assumed for the study. This value guaranteed that the obtained velocity and pressure distributions were not affected by numerical error. The velocity profile from the initial simulation was normalized and adapted for transient time-dependent simulation. The inlet velocity was defined as a user-defined equation that resulted from the product of the extended velocity profile and an input velocity profile based on Doppler measurements. ^23^ The outlet pressure condition was defined on the basis of experimental measurements of blood pressure for a healthy patient. ^24^ The k-omega STT turbulence model was applied. The turbulent kinetic energy (k) and the specific turbulent dispersion coefficient (omega) were also assigned to the studied model.

For the subsequent transient analysis, 200 steps were calculated with a time step of 0.005. A cardiac cycle with a 1 s duration was simulated. The parameters analyzed in the study were velocity and WSS values. The shear stress determines the intensity of the shear forces acting on the vessel surface as a result of fluid flow as follows:

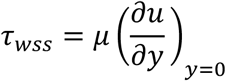

where *μ* is the dynamic viscosity, *u* is the flow velocity parallel to the wall, and *y* is the distance to the wall.

An additional parameter that was analyzed for the selected case was the wall shear stress divergence (WSSD). The wall shear stress divergence considers both the direction and gradient of the WSS, and in regions with high WSSD values, an increased probability of aneurysm occurrence was noted. ^25^

In the simulations, the following geometrical cases were investigated: three cylinder-shaped strings (A – diameter of the string at the widest point of 0.2 mm, which corresponds to a percentage of the string in the cross-sectional area equal to 5.3% of the artery lumen, B – diameter of 0.4 mm, cross-section of 10.6%; C – diameter of 0.56 mm, cross-section of 15%), and two barrel-shaped strings (D – diameter of 0.85 mm, cross-section of 15%; E – diameter of 1.7 mm, cross-section of 30%). The diameters of the strings were chosen considering our results as well as those of previous reports.

## 3. Results

### 3.1. Endovascular anatomy

In 8 out of 30 cases (26.7%) endoscopic assessment revealed the presence of intraluminal structures in the basilar artery (Video 1). Two of these structures were in proximity to the BBA (above the origin of the superior cerebellar arteries), five were near the UVA (below the origin of the anterior inferior cerebellar arteries), and one was in the middle section of the BA (between the origin of the superior and anterior inferior cerebellar arteries). Based on their shape and length, the structures were divided into three groups: strings, septa and chords (Fig. 2 and Fig. 3). The strings (n = 6, 20%) were structures with a length, nearing the diameter of the artery, that was significantly greater than the breadth and depth, which were comparable. The chords (n = 1, 3.33%) were similar but shorter structures, constituting a chord of the diameter of an artery. The septae (n = 1, 3.33%) were structures with a depth and length much greater than their breadth. In each case, intraluminal structures were present in the arteries singularly. Two of the strings were slightly broadened; in one of the cases, the enlargement was balloon shaped, and in the other, it was fusiform shaped (Fig. 3). Interestingly, the strings seemed to be narrowed at their insertion points. The width of the intraluminal structures ranged from 0.1 to 0.5 mm, with a mean value of 0.3 mm, not including two broadened strings, which deformed during fixation and processing and their width was estimated from the angioscopy images (0.7 mm and 1.05 mm).

**Figure 2.**
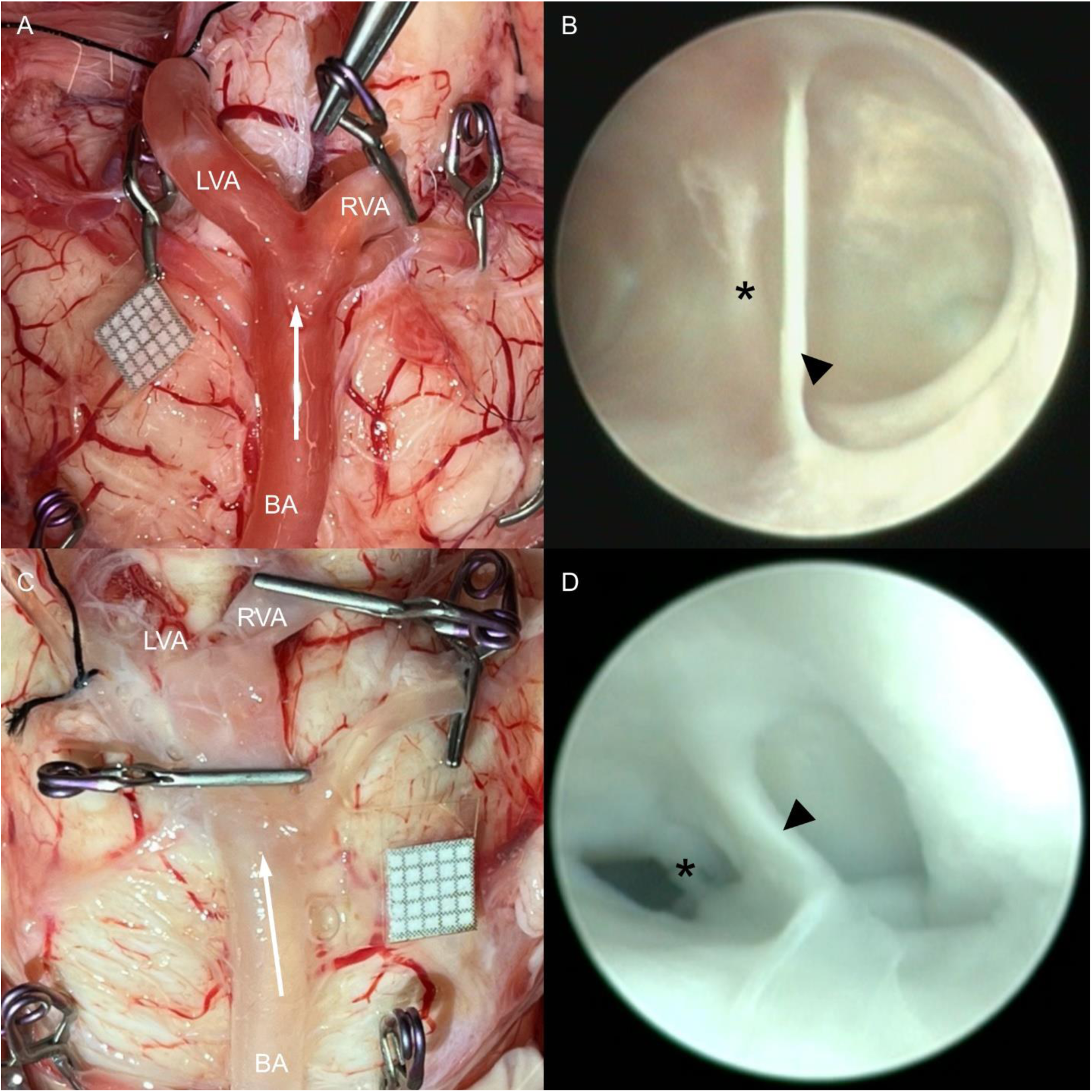
Angioscopy of the basilar artery (BA). **A and C.** Anatomical specimens of the brainstem and corresponding endovascular structures of the basilar artery (**B and D**). The arrows show the direction of introduction of the endoscope. A string (B) and a septum (D) are visible inside the BA (arrowheads) in close proximity to the union of the vertebral arteries (asterisks). BA basilar artery, LVA and RVA left and right vertebral artery.

**Figure 3.**
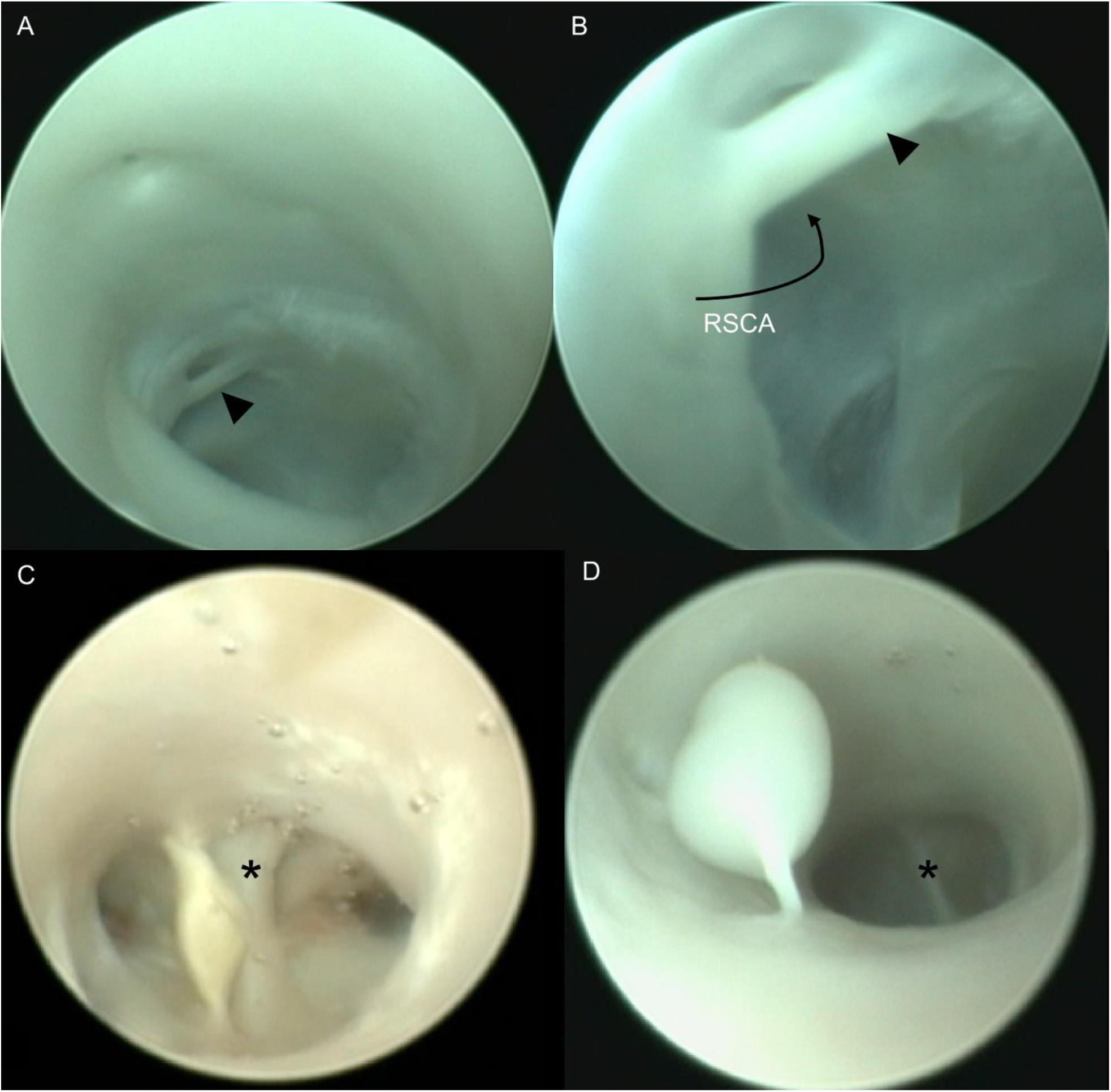
An intravascular chord and atypical (broadened) intravascular strings. **A.** Endoscopic view of a chord (arrowhead) located at the orifice of the right superior cerebellar artery (RSCA). The endoscope was introduced in an opposite direction to that in Fig. 2, i.e., toward the bifurcation of BA. **B.** Zoom of the chord. **C and D.** Endoscopic views of the atypical, broadened strings (arrowheads). The vertebrobasilar junction is marked with an asterisk. RSCA right superior cerebellar artery.

Endoscopy has also revealed several other features of the arterial wall. One of them was the folds present on the arterial walls, which could be divided into two major types: longitudinal (finer, running along the longitudinal axis of the artery) and circular (broader, arranged perpendicularly with respect to the longitudinal axis of the vessel, thicker, spanning the whole circumference of the arterial wall; Supplementary Figure 1 B and C). The folds were visible in all the arteries examined; however, they could be noticed more readily in some cases. Another feature of the arterial wall was translucency, which was prominent in and around arterial bifurcations. The wall of the arterial trunks, conversely, was not translucent (Supplementary Figure 1 A). We did not find any objective measure to assess this aspect; however, both observers (MS and RR) confirmed that they noticed differences between the walls of bifurcations and trunks. Notably, during all of the examinations, the manipulations performed with the endoscope and its occasional prolonged contact with the arterial wall caused severe exfoliation of the endothelium. Moreover, during endoscopic examination, one of the strings detached from one of the insertion points, whereas another one detached completely from the attachments to the arterial wall.

In 18 out of 30 cases (60%), atherosclerosis in the vertebrobasilar system was visualized. The basilar artery was most commonly involved (17 cases, 57%), followed by the left internal carotid artery (13 cases, 37%) and the left middle cerebral artery (11 cases, 33%). Among the 18 cases, 10 were classified as atheromas, 5 as fibroatheromas, and 3 as complicated lesions. Atypical dilation of the basilar artery and stenosis of the right vertebral artery were noted in one case each (3,33%).

### 3.2. Results of the histological examination

Histological evaluation revealed a layered internal structure of intraluminal structures of the basilar artery (Fig. 4). The innermost layer comprised cells morphologically akin to the cells of the adventitia, bordering on more externally located muscular fibers, the internal elastic lamina (readily visualized via Mallory trichrome stain) and the intima. The outermost layer was continuous with the arteries’ intima and endothelium. Although their continuity or thickness might varied across cases, the arrangement of the layers was preserved in all of the structures, regardless of their morphology or site of occurrence. The internal structure of the broadened strings was similar, but the thickness of the layers was higher (Fig. 4 E). In some cases, the arterial wall exhibited thickening of the intima (Fig. 4 A and D). With increased remodeling of the tunica intima, the string attachment sites presented a disrupted morphology, with an altered architecture of the tunica media and a disrupted internal elastic lamina (Fig. 4 A).

**Figure 4.**
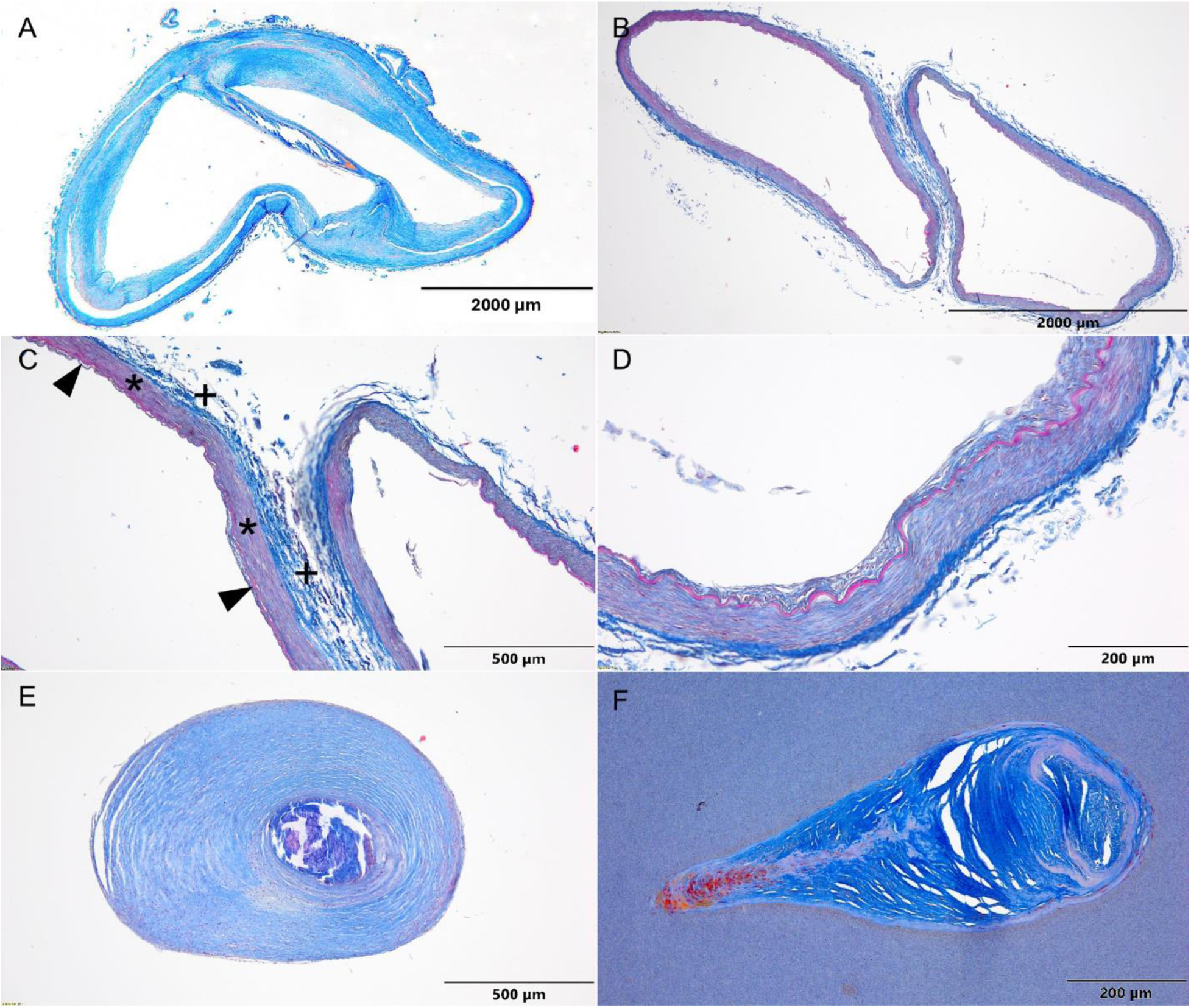
Representative results of the histological studies (Mallory’s trichrome staining). **A and B.** Cross-sections of the basilar arteries and a string (A) or a septum (B) inside. **C.** Zoom of attachment point of the septum. Note the continuity of the tunica intima (arrowheads, the internal elastic lamina), the tunica media (asterisks) and the adventitia (plus sign). **D.** Zoom of intimal hyperplasia of the arterial wall of BA with the septum. **E.** Cross-section of the broadened string shown in Fig. 3D. A layered structure similar to normal strings can be observed, but thickness of the tunica media and adventitia is greater, and the internal structure is partially disrupted. **F.** Cross-section of an asymmetrical string found coincidentally during research on branching points of the pontine arteries. ^26^

### 3.3. Hemodynamic simulation results

The blood flow simulations made it possible to analyze the hemodynamic conditions exerted by the strings. The effect of its presence was greater than that of the local disturbances around the string, but it affected the overall hemodynamic stability of the flow and divided the blood stream.

In all the models, an increase in the average blood flow velocity at the sides of the string and a decrease directly behind it were visualized (Videos 2 and 3), and the severity of the velocity changes and the size of the perturbation areas depended on the string diameter (Fig. 5, left column). Wider strings caused more significant velocity changes—changes in the volume of the blood with increased velocity up to 0.4 m/s were up to 20 times greater in case of the 1.7 mm string (Fig. 5 E) than in the case of the 0.2 mm string (Fig. 5 A). The distribution of time-averaged wall shear stresses (WSS) also showed a repetitive pattern: there were local minima on the front (the velocity decreases after blood impacts the string) and rear surfaces and local maxima on its sides (the higher gradient of the velocity), whereas the WSS increased on the vessel wall surface at the level of the string, excluding areas around the string attachments, where there was a complex pattern of local minima and maxima (Videos 4 and 5, Fig. 5). Behind the strings, there were areas of recirculation, as confirmed by WSSD analysis (Supplementary Figure 2). The wall shear stress disturbances were most severe on the vessel wall around wider strings and on the lateral surfaces of narrow strings, where the stresses reached extremely high values (up to 52 Pa).

**Figure 5.**
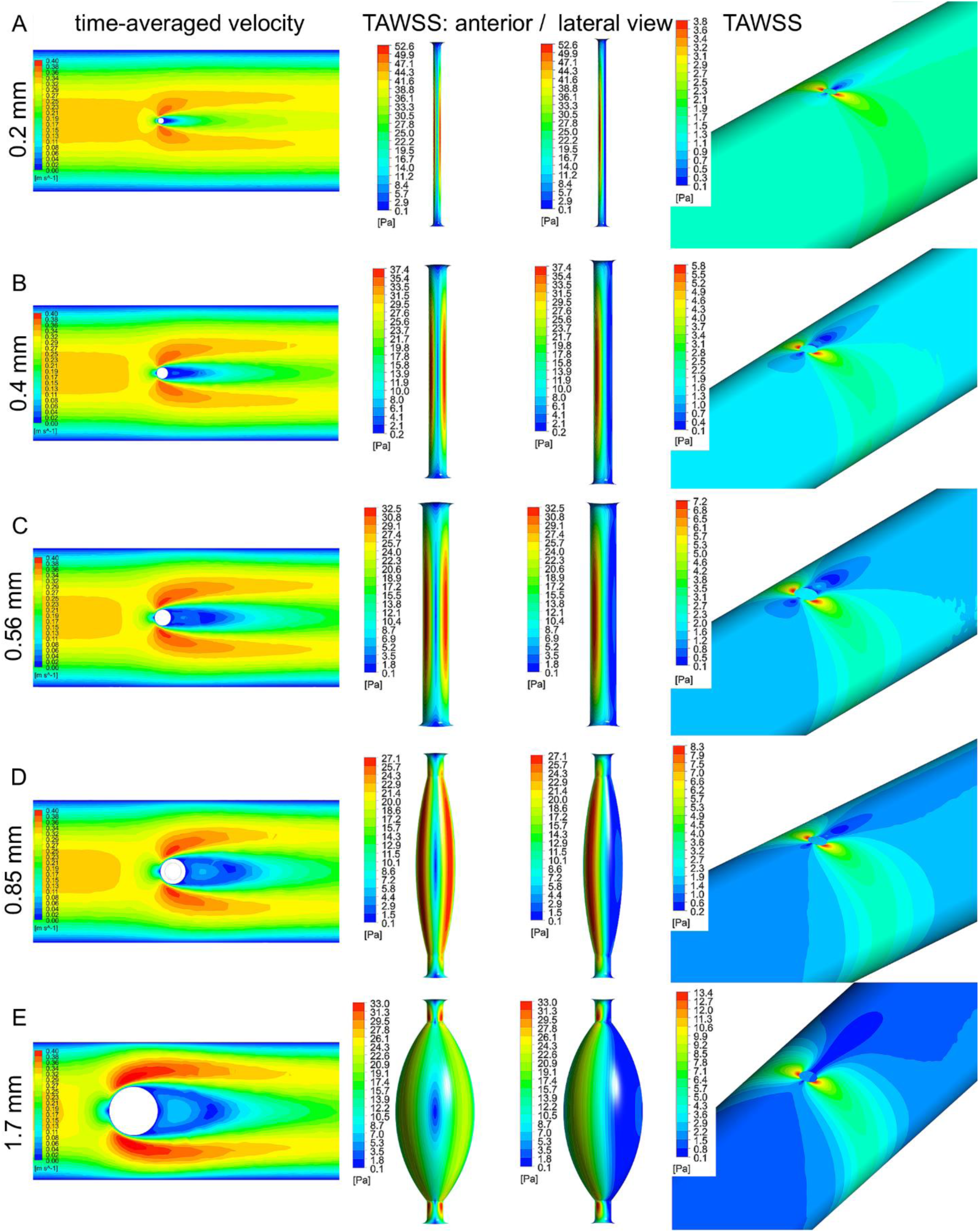
Results of the blood flow simulations: maps of time-averaged velocity (left column), time-averaged wall shear stress of the strings (TAWSS, middle column, anterior and lateral views), time-averaged wall shear stress of the arterial wall (TAWSS, right column). Blood flow velocity increases lateral to and decreases behind the strings. Wall shear stress values reach local maxima on the lateral surfaces of the strings, while TAWSS on the anterior and posterior surfaces is low. The vessel wall is subjected to local TAWSS increase at the level of the string. The severity of hemodynamic changes is related to string diameter, which is indicated by the numbers in the left margin.

## 4. Discussion

### 4.1. Incidence of endovascular structures and study methodology

We present the first and only study that utilized angioscopy to visualize and assess the intraluminal anatomy of basal cerebral arteries (some endoscopic studies concerning the dural sinuses and coronary vessels have been published previously).^19,20,27^ It is also one of a few works focused on this particular matter since 1839, when John Davy first described “peculiarities” in the basilar arteries.^14-16,18,28^ The presence of intraluminal structures in the basal cerebral arteries was also very briefly mentioned by Stehbens.^17^ The methodology of these studies was manifold—two of them were based entirely on anatomical dissection, one focused exclusively on radiological imaging (computed tomography), one on experimental radiological methods (plain radiography and roentgen crystallography), and one included both dissection and histological examination of intraluminal structures. The incidence rates of endovascular entities in basilar arteries reported in earlier studies were 17.4% (Davy), 0.67% (Tubbs et al.), 0.6% (Small et al.), and 10% (Glennon et al.). In the present study, we discovered endovascular structures in 8 out of 30 basilar arteries, accounting for 26.7%, which is the largest value reported in the literature. These results suggested that endoscopy might be a more reliable and sensitive method for assessing endovascular anatomy compared with dissection (with or without an operative microscope). These methods have several drawbacks, which explain the discrepancies between the results.

Classic or microscopic dissection of the basal cerebral arteries is an invasive method that disrupts the original conformation of the vessel, disallowing reliable assessment of the intravascular anatomy. It also may result in unintentional damage to intravascular structures, which may be very delicate as shown in this study. Therefore, some of these structures might be overlooked by researchers. Additionally, if the structures are damaged before dissection, resulting in their detachment from the vessel wall (during acquisition, fixation or decay processes), examination might be falsely negative. Conversely, angioscopy minimizes the risk of damaging the sample and allows for the most credible assessment of endovascular anatomy, as original topographical and anatomical arrangements are preserved, approximated to *in vivo* conditions as much as possible in a postmortem study.

Studies based on imaging modalities also have limitations. Hassler reported that only the largest intravascular structures could by visualized using “ordinary roentgenologic methods”, which at the time comprised plain radiography and angiography.^28^ Small et al. described intraluminal entities in a large group (3509 patients) on the basis of CT angiography.^16^ Surprisingly, the entities corresponding to intraluminal structures found in our study were present in only 21 cases (0.6%), representing a reduction by approximately 50 times. Similar to imaging the perforating arteries,^29^ the width of intravascular structures is too small to visualize all of them via CT angiography; therefore, studies based solely on imaging methods might not reliably represent the incidence of these entities.

### 4.2. Embryologic origin of the endovascular structures

In our opinion, the intravascular entities described in this study, despite differences in their morphology, share a common embryological origin. Fusion of longitudinal neural arteries (LNA, precursors to adult basilar arteries and parts of vertebral arteries) begins on approximately the 29^th^ day of embryonic life on the ventral surface of the pons, simultaneously on different segments of the arteries.^30^ Until the seventh week of gestation, embryonic cerebral vessels’ walls are comprised of a single endothelial layer – the *tunica media* begins to form during the eighth week, and when the adventitia develops at the end of 20^th^ week, the vessels start to resemble their adult form.^31^ Basilar fenestrations—corresponding to segments of longitudinal neural arteries non-fusion—are therefore remnants of the early stage of organization of the CNS vasculature. As seen in the histological samples, intravascular structures are composed of layers corresponding to layers of the arterial wall. The innermost part of the strings and septa is morphologically akin to the adventitia of the same vessel, which advocates for incomplete side-to-side fusion of two arterial channels. This finding confirmed our hypothesis that septa and strings are formed in the same way as fenestrations. According to Lasjaunias et al., ^32^ this indicates that these are not true anatomical variants but rather persistent, stable results of temporary obstacles that emerged during the embryonic period and segmentally prevented longitudinal neural arteries from fusing. This finding is in accordance with previous histological examinations of similar intraarterial structures.^15^ Moreover, a trough corresponding to the septum was visible on the external surface of the basilar artery, suggesting external morphological resemblance between fenestrations and intraluminal structures.

The uneven occurrence of intraarterial entities along the basilar artery is a particularly interesting question. Our findings and reports from other authors have shown that intraluminal structures are more prevalent in the proximity of the union of vertebral arteries. According to most authors, side-to-side fusion of longitudinal neural arteries occurs craniocaudally.^30,33-39^ Proximal nonfusion of the basilar artery, in the form of fenestrations, is far more common than distal fenestrations.^40^ The temporal differences in the fusion of the segments of the basilar artery seem to account for the disproportionate occurrence of fenestrations along the vertebrobasilar system.^41^ The rationale for this phenomenon could be that the probability of incomplete fusion becomes greater at later stages of cerebrovascular development, as hemodynamic conditions in the vertebrobasilar system become more established with time and more closely resemble the ultimate conformation of cerebral arteries. From an embryological perspective, intraluminal structures can be viewed as abnormalities that are closely related to fenestrations; this rationale would also be applicable to their predilection to occur in the proximal segment of the BA.^30,33-39^

### 4.3. Classification of intraluminal structures

Authors of earlier works on this topic have proposed various classifications of the encountered structures: Hassler proposed simple segregation into two groups: calcified and noncalcified “bridges”; Small et al. proposed distinguishing between fenestrations and aberrant fenestrations of the basilar artery, which could assume different sizes and shapes (funnels, hourglasses) and be accompanied by internal nodules or calcifications; Glennon et al. suggested that the bands should be divided into those that are sagitally oriented (dubbed “vertical” in the original publication) and those that are horizontally oriented.

Considering vascular neuroembryology and possible rotation of the basilar artery, it seems rather improbable that incomplete fusion of LNAs would result exclusively in sagitally oriented bands. Most importantly, all of the entities described in this and previous studies seem to share a common embryological origin. Given this evidence, we propose to desist from classifying intraluminal basilar bands into discrete morphological entities. Instead, we strongly suggest aggregating all the described structures (bands, bridges, aberrant basilar fenestrations, chords, strings, and septa) under one term and conceptualizing them as <u>basilar nonfusion spectrum</u> <u>(BNFS)</u>. The rationale for this outlook is based primarily on the fact that fusion of LNAs is a continuous process, which might terminate at numerous points in time (albeit more frequently at later stages). Therefore, all possible configurations (including fenestrations) fall somewhere between the most commonly observed basilar artery with a single vascular canal and rare cases of complete nonfusion of the basilar artery. These cases constitute two opposite extremes of the spectrum. From an embryological perspective, true fenestration and intraluminal strings are in fact the same phenomenon, with the only difference being their extent (Fig. 6).

**Figure 6.**
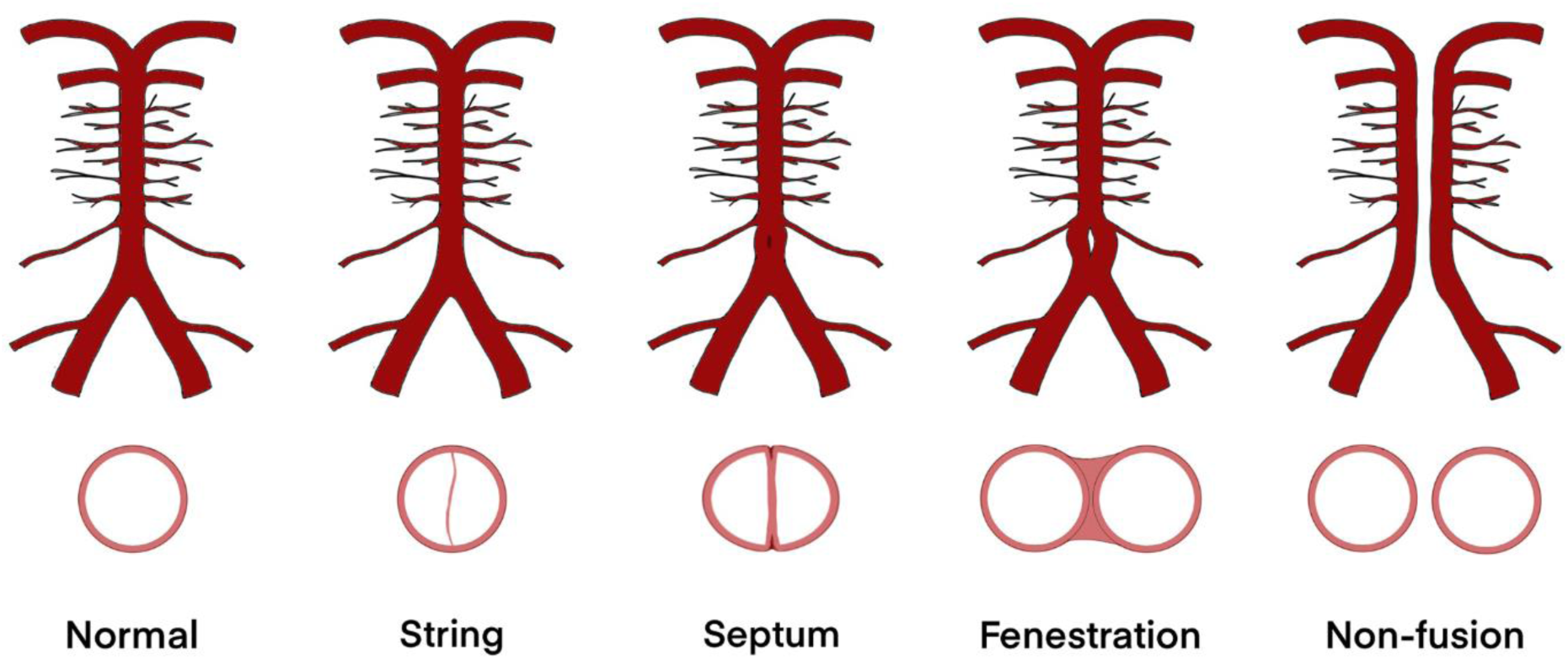
Basilar nonfusion spectrum scheme. Endovascular strings, septa, and fenestrations fall between the most commonly observed basilar artery with a single vascular canal and rare cases of complete nonfusion of the basilar artery, and represent consequences of incomplete embryological fusion of the longitudinal neural arteries.

### 4.4. Clinical significance

The subject of this research is relevant in the clinical context, and hemodynamic consequences should be discussed with respect to the results of the blood flow simulations. The clinical implications are related to three issues: endovascular interventions, predisposition to neurovascular disease, and radiologic imaging.

Currently, being aware of the existence of intraluminal structures in the vertebrobasilar system seems to be more clinically relevant, as intravascular techniques have become an established standard in the treatment of cerebrovascular disorders. Owing to the lack of widespread knowledge of the existence of endovascular structures and suitable *in vivo* diagnostic modalities, the consequences of their presence on the performance of interventions or complications have not yet been described. Yoon et al. reported that basilar fenestrations are not obstacles or contraindications for intravascular management of aneurysms of posterior cerebral circulation;^42^ however, endoluminal structures are not visible in radiological images, as discussed above. Potential intraoperative difficulties, such as physical resistance of unknown origin during stent expansion or catheter manipulations, can be expected. A string may also detach from the vessel wall, occlude a distal branch (as observed in our study during one of the experiments), and cause unexplained ischemic complications. It can be presumed that the endothelium exfoliation areas observed during experiments because of contact with the endoscope may also develop as a result of catheter manipulation. Moreover, the site of arterial damage (string attachment point or exfoliated area) may promote hemostasis and produce thromboembolic material. Considered high prevalence of endovascular structures of the BA, these mechanisms might explain, to some extent, ischemic complications after stenting of the basilar artery or aneurysm coiling (including the highest rate of complications and poor outcome associated with flow diversion of the UVA and basilar trunk), ^43-46^ the reduced effectiveness of endovascular thrombectomy in restoring blood flow in about 30% of patients with basilar artery occlusion, ^47,48^ increased risk of thromboembolic complications after thrombectomy in the posterior circulation, ^49^ as well as technical difficulties.

The blood flow simulations revealed that the presence of intraluminal strings might promote atherosclerosis development and neointimal remodeling of the arterial wall. The distribution of the hemodynamic changes follows a repeatable pattern: areas of elevated WSS are located on the lateral surfaces of the strings and on the vessel wall at the level of the string, whereas areas of low WSS are located behind the attachment points of the strings and on their front and back surfaces. In addition, there are areas of flow stagnation and recirculation behind the string attachment sites. High WSS values are thought to cause the formation of high-risk plaques, ^50^ whereas low WSS values predispose to neointimal thickening and atherosclerosis development.^51-55^ Remodeling of the arterial wall is strongly stimulated by the multidirectional distribution of the WSS, recirculation, and stagnation of flow, ^56,57^ so it can be expected at the level of the string and behind its attachments, where the vessel wall is particularly vulnerable. Indeed, in some cases, we observed neointimal hyperplasia of the basilar artery (Fig. 4 A and D). The strings themselves are also subject to remodeling, as can be seen in cross sections of the broadened ones (Fig. 4 E). Interestingly, we found a similar asymmetrically remodeled string during histological studies of the pontine arteries (Fig. 4 F).^26^

It is worth emphasizing the role of the pontine arteries, the initially narrowed branching points of which ^26^ can thus be remodeled and closed, leading to devastating neurological complications. ^58,59^ Notably, the observed hemodynamic changes stimulate thrombus formation, ^53,55,57^ which may further promote thromboembolic complications, as observed in cases of carotid webs. ^60^ Moreover, a high percentage of basilar artery occlusion cases has undetermined cause. ^61^ There are known isolated cases of ischemic stroke attributed to the presence of uncharacteristic structures within the basilar artery. ^62-66^ The authors of these reports attempted to call the structures by various names (fenestrations, basilar web, “protruding basilar artery lesion”) but emphasized their unusual morphology. Given the above described limitations of the radiological imaging methods, these findings may correspond to intravascular strings or septa.

The relationship between the development of aneurysms and the presence of intravascular structures requires further investigation. The distribution of shear stress suggests the presence of remodeling-prone areas near the attachment sites of the strings, where a locally significantly elevated WSS is observed. ^67^ In our study, the areas were histologically distinct from the rest of the arterial wall and presented discontinuation of the internal elastic lamina and changes in the architecture of the tunica media. In addition to the string attachment sites, WSSD analysis identified an additional area behind the string as susceptible to aneurysm initiation.^25,68^ The situation analyzed may be analogous to arterial fenestration, which predisposes patients to aneurysm development. ^69,70^

Finally, the intraluminal septum found in this study was accompanied by slight dilation of the basilar artery along the structure. In angiography (classic, CT or MRI), such alterations in vessel diameter might be mistaken for aneurysm, dissection, or other cerebrovascular pathology, resulting in misdiagnosis and submission of patients to unnecessary treatment.

### 4.5. Limitations of the study and perspectives

The described method and its results are subject to several limitations. Anatomical specimens from Europeans have been studied, and it is necessary to investigate the prevalence of intravascular structures in other populations. Postmortem studies often do not allow for complete analysis of risk factors, but the material presented was sufficient to meet the purpose of the study, that is, to describe intravascular structures and investigate hemodynamic conditions. The numerical simulations assumed stiff surfaces of the strings and BA; therefore, the WSS values might be overestimated; the boundary conditions were based on the literature. However, it must be emphasized that more important than the absolute value of the WSS is the distribution of areas with relatively high or low WSSs. ^57,71,72^ Perhaps in the near future, improvements in radiological imaging methods will make it possible to visualize intravascular structures in vivo and determine their significance in relation to cerebrovascular disease and endovascular interventions.

## Data Availability

Data supporting the conclusions are presented in the article.

## ACKNOWLEDGEMENTS

Michał Kucewicz and Radosław Rzepliński are recipients of the Foundation for Polish Science scholarship.

## DISCLOCURES

None.

## SUPPLEMENTARY FILE CAPTIONS

**Supplementary** Figure 1 Angioscopy of the basilar artery. **A.** Translucent areas at the branching points (asterisks). **B and C.** Longitudinal (B) and circular (C) folds of the endothelium.

**Supplementary** Figure 2 Distribution of the wall shear stress divergence (WSSD). Areas of elevated WSSD are localized on the front and rear surfaces of the string, on the string attachment sites, and behind the string on the arterial wall. The last area corresponds to recirculation and reattachment site of the blood flow.

**Video 1** Angioscopy of the basilar artery. A string is visible in close proximity to the union of the vertebral arteries.

**Videos 2-5** Results of the blood flow simulations. Animations of blood flow velocity distribution: longitudinal (Video 2) and cross-section (Video 3) through the basilar artery. Animations of the wall shear stress distribution: on the string surface (Video 4) and on the arterial wall (Video 5). Blood flow direction is from left to right. Animations present one cardiac cycle.

## REFERENCES

1. Alemseged F, Nguyen TN, Alverne FM, Liu X, Schonewille WJ, Nogueira RG. Endovascular Therapy for Basilar Artery Occlusion. Stroke. 2023;54:1127–1137. doi: 10.1161/STROKEAHA.122.040807

2. Elder TA, White TG, Woo HH, Siddiqui AH, Nunna R, Siddiq F, Esposito G, Chang D, Gonzalez NR, Amin-Hanjani S. Future of Endovascular and Surgical Treatments of Atherosclerotic Intracranial Stenosis. Stroke. 2024;55:344–354. doi: 10.1161/STROKEAHA.123.043634

3. Ospel JM, Nguyen TN, Jadhav AP, Psychogios MN, Clarencon F, Yan B, Goyal M. Endovascular Treatment of Medium Vessel Occlusion Stroke. Stroke. 2024;55:769–778. doi: 10.1161/STROKEAHA.123.036942

4. Tjoumakaris SI, Hanel R, Mocco J, Ali-Aziz Sultan M, Froehler M, Lieber BB, Coon A, Tateshima S, Altschul DJ, Narayanan S, et al. ARISE I Consensus Review on the Management of Intracranial Aneurysms. Stroke. 2024;55:1428–1437. doi: 10.1161/STROKEAHA.123.046208

5. Kan P, Fiorella D, Dabus G, Samaniego EA, Lanzino G, Siddiqui AH, Chen H, Khalessi AA, Pereira VM, Fifi JT, et al. ARISE I Consensus Statement on the Management of Chronic Subdural Hematoma. Stroke. 2024;55:1438–1448. doi: 10.1161/STROKEAHA.123.044129

6. Lasjaunias P, Brugge K, Berenstein A. Embryological and Anatomical Introduction. In: Lasjaunias P, Brugge K, Berenstein A, eds. Surgical Neuroangiography. Berlin: Springer; 2006:1-25.

7. Lasjaunias PL, Berenstein A, Brugge KGt. Surgical neuroangiography. 2nd ed. Berlin; New York: Springer; 2001.

8. Marinkovic S, Gibo H, Milisavljevic M, Cetkovic M. Anatomic and clinical correlations of the lenticulostriate arteries. Clin Anat. 2001;14:190–195. doi: 10.1002/ca.1032

9. Marinkovic SV, Gibo H. The surgical anatomy of the perforating branches of the basilar artery. Neurosurgery. 1993;33:80–87. doi: 10.1227/00006123-199307000-00012

10. Marinkovic SV, Kovacevic MS, Marinkovic JM. Perforating branches of the middle cerebral artery. Microsurgical anatomy of their extracerebral segments. J Neurosurg. 1985;63:266–271. doi: 10.3171/jns.1985.63.2.0266

11. Rhoton AL, Jr. The cerebellar arteries. Neurosurgery. 2000;47:S29–68. doi: 10.1097/00006123-200009001-00010

12. Rhoton AL, Jr. The supratentorial arteries. Neurosurgery. 2002;51:S53–120.

13. Rzeplinski R, Tomaszewski M, Slugocki M, Karczewski K, Krajewski P, Skadorwa T, Malachowski J, Ciszek B. Method of creating 3D models of small caliber cerebral arteries basing on anatomical specimens. J Biomech. 2021;125:110590. doi: 10.1016/j.jbiomech.2021.110590

14. Davy J. Of a Peculiarity of Structure Occasionally Occurring in the Basilar Artery of Man.Edinb Med Surg J. 1839;51:70–75.

15. Glennon SE, Ram K, Gupta T, Iwanaga J, Dumont AS, Small JE, Sahni D, Tubbs RS. Basilar Artery Bands: Anatomic and Histologic Study with Application to Coiling and Stenting Procedures. World Neurosurg. 2022;160:e227–e233. doi: 10.1016/j.wneu.2021.12.114

16. Small JE, Macey MB, Wakhloo AK, Sehgal S. CTA Evaluation of Basilar Septations: An Entity Better Characterized as Aberrant Basilar Fenestrations. AJNR Am J Neuroradiol. 2021;42:701–707. doi: 10.3174/ajnr.A7008

17. Stehbens WE. Flow in experimental models simulating intravascular cords traversing the arterial lumen. Vasc Surg. 1975;9:132–140. doi: 10.1177/153857447500900302

18. Tubbs RS, Shaffer WA, Loukas M, Shoja MM, Harrigan MR, Oakes WJ. Intraluminal septation of the basilar artery: incidence and potential clinical significance. Folia Morphol (Warsz). 2008;67:193–195.

19. Sharifi M, Kunicki J, Krajewski P, Ciszek B. Endoscopic anatomy of the chordae willisii in the superior sagittal sinus. J Neurosurg. 2004;101:832–835. doi: 10.3171/jns.2004.101.5.0832

20. Zawadzki M, Pietrasik A, Pietrasik K, Marchel M, Ciszek B. Endoscopic study of the morphology of Vieussens valve. Clin Anat. 2004;17:318–321. doi: 10.1002/ca.10229

21. Tomaszewski M, Kucewicz M, Rzepliński R, Małachowski J, Ciszek B. Numerical aspects of modeling flow through the cerebral artery system with multiple small perforators. Biocybernetics and Biomedical Engineering. 2024;44:341–357. doi: 10.1016/j.bbe.2024.04.002

22. Tomaszewski M, Sybilski K, Baranowski P, Małachowski J. Experimental and numerical flow analysis through arteries with stent using particle image velocimetry and computational fluid dynamics method. Biocybernetics and Biomedical Engineering. 2020;40:740–751.

23. Deeg KH, Reisig A. Doppler sonographic screening of the flow in the basilar artery during head rotation reduces the risk for sudden infant death. Ultraschall Med. 2010;31:506–514. doi: 10.1055/s-0029-1245144

24. Blanco PJ, Muller LO, Spence JD. Blood pressure gradients in cerebral arteries: a clue to pathogenesis of cerebral small vessel disease. Stroke Vasc Neurol. 2017;2:108–117. doi: 10.1136/svn-2017-000087

25. Tanaka K, Takao H, Suzuki T, Fujimura S, Uchiyama Y, Otani K, Ishibashi T, Mamori H, Fukudome K, Yamamoto M, et al. Relationship between hemodynamic parameters and cerebral aneurysm initiation. Annu Int Conf IEEE Eng Med Biol Soc. 2018;2018:1347–1350. doi: 10.1109/EMBC.2018.8512466

26. Rzeplinski R, Tarka S, Tomaszewski M, Kucewicz M, Acewicz A, Małachowski J, Ciszek B. Narrowings of the deep cerebral perforating arteries ostia: geometry, structure, and clinical implications. Journal of Stroke. 2024;In Press.

27. Ye Y, Ding J, Huang S, Wang Q. Related Structures in the Straight Sinus: An Endoscopic Anatomy and Histological Study. Front Neuroanat. 2020;14:573217. doi: 10.3389/fnana.2020.573217

28. Hassler O. Intra-Arterial Bridges in the Larger Cerebral Arteries. Acta Radiol Diagn (Stockh). 1965;3:305–309. doi: 10.1177/028418516500300403

29. Rzeplinski R, Slugocki M, Kwiatkowska M, Tarka S, Tomaszewski M, Kucewicz M, Karczewski K, Krajewski P, Malachowski J, Ciszek B. Standard clinical computed tomography fails to precisely visualise presence, course and branching points of deep cerebral perforators. Folia Morphol (Warsz). 2023;82:37–41. doi: 10.5603/FM.a2021.0133

30. Padget D. The development of the cranial arteries in the human embryo. Washington; 1948.

31. Fujimoto K. ’Medial defects’ in the prenatal human cerebral arteries: an electron microscopic study. Stroke. 1996;27:706–708. doi: 10.1161/01.str.27.4.706

32. Lasjaunias P, Berenstein A, Brugge KG. Clinical Vascular Anatomy and Variations. Berlin, Heidelberg: Springer Berlin, Heidelberg; 2001.

33. Finlay HM, Canham PB. The layered fabric of cerebral artery fenestrations. Stroke. 1994;25:1799–1806. doi: 10.1161/01.str.25.9.1799

34. Goldstein JH, Woodcock R, Do HM, Phillips CD, Dion JE. Complete duplication or extreme fenestration of the basilar artery. AJNR Am J Neuroradiol. 1999;20:149–150.

35. Raybaud C. Normal and abnormal embryology and development of the intracranial vascular system. Neurosurg Clin N Am. 2010;21:399–426. doi: 10.1016/j.nec.2010.03.011

36. Shroff M, Blaser S, Jay V, Chitayat D, Armstrong D. Basilar artery duplication associated with pituitary duplication: a new finding. AJNR Am J Neuroradiol. 2003;24:956–961.

37. Uchino A, Sawada A, Takase Y, Fujita I, Kudo S. Extreme fenestration of the basilar artery associated with cleft palate, nasopharyngeal mature teratoma, and hypophyseal duplication. Eur Radiol. 2002;12:2087–2090. doi: 10.1007/s00330-001-1194-0

38. Hoh BL, Rabinov JD, Pryor JC, Hirsch JA, Dooling EC, Ogilvy CS. Persistent nonfused segments of the basilar artery: longitudinal versus axial nonfusion. AJNR Am J Neuroradiol. 2004;25:1194–1196.

39. Krings T, Baccin CE, Alvarez H, Ozanne A, Stracke P, Lasjaunias PL. Segmental unfused basilar artery with kissing aneurysms: report of three cases and literature review. Acta Neurochir (Wien). 2007;149:567–574; discussion 574. doi: 10.1007/s00701-007-1118-0

40. Tran-Dinh HD, Soo YS, Jayasinghe LS. Duplication of the vertebro-basilar system. Australas Radiol. 1991;35:220–224. doi: 10.1111/j.1440-1673.1991.tb03012.x

41. Tasker AD, Byrne JV. Basilar artery fenestration in association with aneurysms of the posterior cerebral circulation. Neuroradiology. 1997;39:185–189. doi: 10.1007/s002340050389

42. Yoon SM, Chun YI, Kwon Y, Kwun BD. Vertebrobasilar junction aneurysms associated with fenestration: experience of five cases treated with Guglielmi detachable coils. Surg Neurol. 2004;61:248–254. doi: 10.1016/S0090-3019(03)00485-3

43. Adeeb N, Griessenauer CJ, Dmytriw AA, Shallwani H, Gupta R, Foreman PM, Shakir H, Moore J, Limbucci N, Mangiafico S, et al. Risk of Branch Occlusion and Ischemic Complications with the Pipeline Embolization Device in the Treatment of Posterior Circulation Aneurysms. AJNR Am J Neuroradiol. 2018;39:1303–1309. doi: 10.3174/ajnr.A5696

44. Davis MC, Deveikis JP, Harrigan MR. Clinical Presentation, Imaging, and Management of Complications due to Neurointerventional Procedures. Semin Intervent Radiol. 2015;32:98–107. doi: 10.1055/s-0035-1549374

45. Alwakeal A, Shlobin NA, Golnari P, Metcalf-Doetsch W, Nazari P, Ansari SA, Hurley MC, Cantrell DR, Shaibani A, Jahromi BS, et al. Flow Diversion of Posterior Circulation Aneurysms: Systematic Review of Disaggregated Individual Patient Data. AJNR Am J Neuroradiol. 2021;42:1827–1833. doi: 10.3174/ajnr.A7220

46. Wang CB, Shi WW, Zhang GX, Lu HC, Ma J. Flow diverter treatment of posterior circulation aneurysms. A meta-analysis. Neuroradiology. 2016;58:391–400. doi: 10.1007/s00234-016-1649-2

47. Liu X, Dai Q, Ye R, Zi W, Liu Y, Wang H, Zhu W, Ma M, Yin Q, Li M, et al. Endovascular treatment versus standard medical treatment for vertebrobasilar artery occlusion (BEST): an open-label, randomised controlled trial. Lancet Neurol. 2020;19:115–122. doi: 10.1016/S1474-4422(19)30395-3

48. Langezaal LCM, van der Hoeven E, Mont’Alverne FJA, de Carvalho JJF, Lima FO, Dippel DWJ, van der Lugt A, Lo RTH, Boiten J, Lycklama ANGJ, et al. Endovascular Therapy for Stroke Due to Basilar-Artery Occlusion. N Engl J Med. 2021;384:1910–1920. doi: 10.1056/NEJMoa2030297

49. Yeo LLL, Holmberg A, Mpotsaris A, Soderman M, Holmin S, Kuntze Soderqvist A, Ohlsson M, Bhogal P, Gontu V, Andersson T, et al. Posterior Circulation Occlusions May Be Associated with Distal Emboli During Thrombectomy : Factors for Distal Embolization and a Review of the Literature. Clin Neuroradiol. 2019;29:425–433. doi: 10.1007/s00062-018-0679-z

50. Eshtehardi P, Brown AJ, Bhargava A, Costopoulos C, Hung OY, Corban MT, Hosseini H, Gogas BD, Giddens DP, Samady H. High wall shear stress and high-risk plaque: an emerging concept. Int J Cardiovasc Imaging. 2017;33:1089–1099. doi: 10.1007/s10554-016-1055-1

51. Zhou M, Yu Y, Chen R, Liu X, Hu Y, Ma Z, Gao L, Jian W, Wang L. Wall shear stress and its role in atherosclerosis. Front Cardiovasc Med. 2023;10:1083547. doi: 10.3389/fcvm.2023.1083547

52. Malik J, L N, A V, E C, V L, K BS, L L, T G, M P, P M. Wall Shear Stress Alteration: a Local Risk Factor of Atherosclerosis. Curr Atheroscler Rep. 2022;24:143–151. doi: 10.1007/s11883-022-00993-0

53. Urschel K, Tauchi M, Achenbach S, Dietel B. Investigation of Wall Shear Stress in Cardiovascular Research and in Clinical Practice-From Bench to Bedside. Int J Mol Sci. 2021;22. doi: 10.3390/ijms22115635

54. Wang X, Shen Y, Shang M, Liu X, Munn LL. Endothelial mechanobiology in atherosclerosis. Cardiovasc Res. 2023;119:1656–1675. doi: 10.1093/cvr/cvad076

55. Ng J, Bourantas CV, Torii R, Ang HY, Tenekecioglu E, Serruys PW, Foin N. Local Hemodynamic Forces After Stenting: Implications on Restenosis and Thrombosis. Arterioscler Thromb Vasc Biol. 2017;37:2231–2242. doi: 10.1161/ATVBAHA.117.309728

56. Weinberg PD. Haemodynamic Wall Shear Stress, Endothelial Permeability and Atherosclerosis-A Triad of Controversy. Front Bioeng Biotechnol. 2022;10:836680. doi: 10.3389/fbioe.2022.836680

57. Chiu JJ, Chien S. Effects of disturbed flow on vascular endothelium: pathophysiological basis and clinical perspectives. Physiol Rev. 2011;91:327–387. doi: 10.1152/physrev.00047.2009

58. Kwiatkowska M, Rzeplinski R, Ciszek B. Anatomy of the pontine arteries and perforators of the basilar artery in humans. J Anat. 2023;243:997–1006. doi: 10.1111/joa.13927

59. Zheng J, Sun B, Lin R, Teng Y, Zheng E, Zhao X, Xue Y. Basilar artery plaque distribution is associated with pontine infarction and vertebrobasilar artery geometry. Front Neurol. 2023;14:1079905. doi: 10.3389/fneur.2023.1079905

60. Mei J, Chen D, Esenwa C, Gold M, Burns J, Zampolin R, Slasky SE. Carotid web prevalence in a large hospital-based cohort and its association with ischemic stroke. Clin Anat. 2021;34:867–871. doi: 10.1002/ca.23735

61. Alemseged F, Nguyen TN, Coutts SB, Cordonnier C, Schonewille WJ, Campbell BCV. Endovascular thrombectomy for basilar artery occlusion: translating research findings into clinical practice. Lancet Neurol. 2023;22:330–337. doi: 10.1016/S1474-4422(22)00483-5

62. Li J, Chen L. Brainstem Infarction Due to a Basilar Arterial Web. Radiology. 2023;309:e231106. doi: 10.1148/radiol.231106

63. Esenwa C, Labovitz D, Caplan LR. "Basilar Web" Causing Basilar Branch Infarction. J Stroke Cerebrovasc Dis. 2019;28:104366. doi: 10.1016/j.jstrokecerebrovasdis.2019.104366

64. Fernandez-Vidal JM, Guasch-Jimenez M, Ruiz-Barrio I, Gomez-Anson B, Tecame M, Marti-Fabregas J. Basilar web and basilar fenestration: a case report. Neurologia (Engl Ed). 2024;39:209–210. doi: 10.1016/j.nrleng.2024.01.010

65. Green DM, Caplan LR. Recurrent basilar branch infarcts due to a protruding basilar artery lesion. Cerebrovasc Dis. 2000;10:489. doi: 10.1159/000016118

66. Scherer A, Siebler M, Aulich A. Virtual arterial endoscopy as a diagnostic aid in a patient with basilar artery fenestration and thromboembolic pontine infarct. AJNR Am J Neuroradiol. 2002;23:1237–1239.

67. Meng H, Tutino VM, Xiang J, Siddiqui A. High WSS or low WSS? Complex interactions of hemodynamics with intracranial aneurysm initiation, growth, and rupture: toward a unifying hypothesis. AJNR Am J Neuroradiol. 2014;35:1254–1262. doi: 10.3174/ajnr.A3558

68. Fujimura S, Tanaka K, Takao H, Okudaira T, Koseki H, Hasebe A, Suzuki T, Uchiyama Y, Ishibashi T, Otani K, et al. Computational fluid dynamic analysis of the initiation of cerebral aneurysms. J Neurosurg. 2022;137:335–343. doi: 10.3171/2021.8.JNS211452

69. Black SP, Ansbacher LE. Saccular aneurysm associated with segmental duplication of the basilar artery. A morphological study. J Neurosurg. 1984;61:1005–1008. doi: 10.3171/jns.1984.61.6.1005

70. Crompton MR. The pathology of ruptured middle-cerebral aneurysms with special reference to the differences between the sexes. Lancet. 1962;2:421–425. doi: 10.1016/s0140-6736(62)90281-7

71. Pohl U, Holtz J, Busse R, Bassenge E. Crucial role of endothelium in the vasodilator response to increased flow in vivo. Hypertension. 1986;8:37–44. doi: 10.1161/01.hyp.8.1.37

72. Ward MR, Pasterkamp G, Yeung AC, Borst C. Arterial remodeling. Mechanisms and clinical implications. Circulation. 2000;102:1186–1191. doi: 10.1161/01.cir.102.10.1186

